# Variation in Telehealth Use in a National Home Test-to-Treat Program for Acute Respiratory Infections

**DOI:** 10.64898/2026.05.24.26353984

**Authors:** Wojciech R. Losos, Biqi Wang, Kim Fisher, Laurel O’Connor, Apurv Soni, Ben S. Gerber

**Author notes:** **Corresponding Author:** Wojciech Losos, MD, Program in Digital Medicine, University of Massachusetts Chan Medical School, 55 Lake Avenue North, Worcester, MA 01655.

## Abstract

**Background:** Home Test-to-Treat (HTTT) programs deliver timely antiviral treatment for acute respiratory infections, including COVID-19 and influenza, through at-home testing and telehealth. Because access is often measured by visit occurrence, variation in how and when care is delivered may be overlooked. We hypothesized that telehealth access follows distinct process-based patterns.

**Methods:** We analyzed de-identified encounters from the national HTTT program (September 2023–July 2024); 6,213 of 8,160 eligible individuals remained after exclusions for missing data. Phenotypes were derived by k-means clustering of standardized variables capturing encounter timing, modality preference, process duration, and sociodemographic and digital access attributes. Ten-day surveys assessed symptom duration and healthcare utilization.

**Results:** Three phenotypes emerged: Delayed/Disrupted Access (n = 1,537; 24.7%), Digitally Engaged but Socioeconomically Vulnerable (n = 1,460; 23.5%), and Mainstream Access and Efficient Utilization (n = 3,216; 51.8%). Mean process duration differed (15.93 [SD 3.84] vs 3.69 [3.31] vs 2.87 [2.41] hours; p < 0.001). Synchronous preference was lowest in the Digitally Engaged group (22.9%); antiviral prescribing was high (88.6%–91.9%). Among 10-day respondents (n = 1,023), symptom duration did not differ. Emergency department visits were most frequent in the Digitally Engaged group (2.3% vs 0.0% and 0.5%; p = 0.02) and urgent care in the Delayed/Disrupted group (5.8% vs 4.1% vs 2.0%; p = 0.02).

**Conclusions:** Telehealth use in a national HTTT program formed distinct phenotypes defined by timing, modality, and care-process efficiency. Evaluating equity requires attention to how and when care is delivered, not simply whether it occurred.

## Introduction

Home Test-to-Treat (HTTT) is a federally supported telehealth care model that integrates at-home diagnostic testing, digital result submission, and remote clinical evaluation to enable timely antiviral treatment of acute respiratory infections, including COVID-19 and influenza.^1–3^ The program was designed to shorten the interval between diagnosis and treatment while mitigating common barriers to care, including geographic distance, scheduling constraints, and insurance coverage gaps.^2,4,5^ Early national implementation demonstrated substantial reach and high utilization during evenings and weekends, underscoring telehealth’s expanding role as a primary access pathway for time-sensitive conditions.^1,6,7^

Prior telehealth research has largely focused on comparisons with in-person care, overall utilization, effectiveness, patient satisfaction, and disparities in access.^8–11^ These approaches have provided important insights, but measures based on visit occurrence may not capture clinically meaningful variation in how and when telehealth care is delivered. For acute infections, variation in telehealth delivery may introduce delays at multiple stages of the care process, including the intervals between diagnostic confirmation, clinical evaluation, and treatment initiation.^4^ Even when appropriate treatment is ultimately received, operational delays may contribute to inefficiency, higher downstream utilization, and poorer patient experience, particularly for individuals facing time scarcity.^12,13^ Telehealth availability alone may therefore mask process-level inequities, in which care is technically accessible but delayed, fragmented, or inefficient.^14–16^

The nationwide HTTT program—which provides free COVID-19 and influenza testing and treatment via telehealth—offers a unique opportunity to examine heterogeneity in telehealth access beyond visit completion. We sought to identify telehealth access phenotypes based on HTTT encounter timing, modality, and care-process efficiency, and to examine how these phenotypes differed with respect to sociodemographic characteristics, telehealth delivery, and subsequent healthcare utilization. We conceptualized telehealth access as a process shaped by timing, modality, and care-process efficiency, rather than simply by whether a visit occurred (Figure 1).

**Figure 1.**
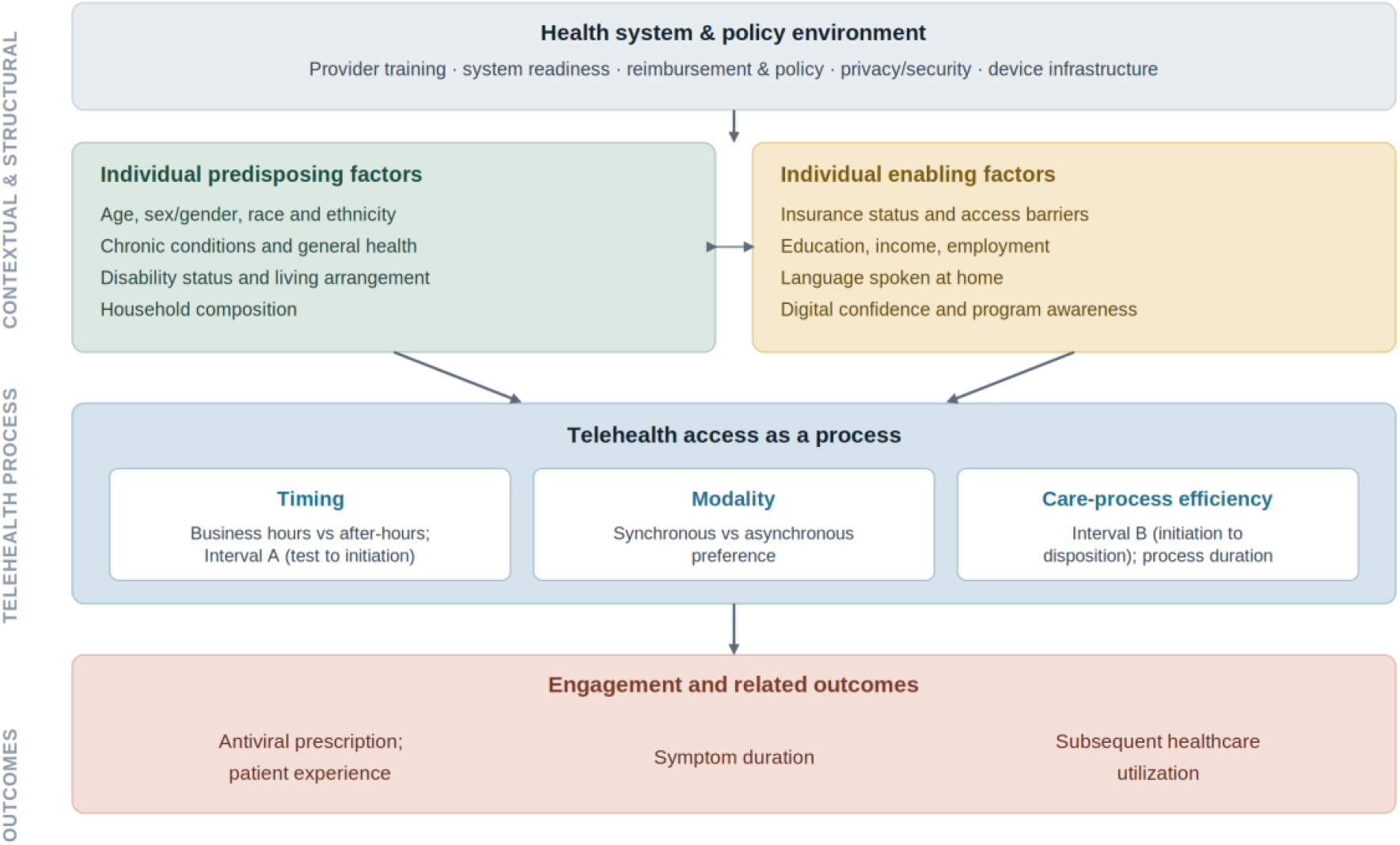
Conceptual framework of telehealth access as a process. Telehealth access is shaped by contextual and structural determinants—the health system and policy environment, individual predisposing factors, and individual enabling factors—and is expressed through telehealth process variables: timing (including Interval A), modality preference, and care-process efficiency (including Interval B). These processes, in turn, relate to engagement and downstream outcomes, including antiviral prescription, symptom duration, patient experience, and subsequent healthcare utilization. The framework emphasizes that access is a process rather than a single event defined by visit occurrence.

## Methods

### Study Design and Setting

We conducted a retrospective cohort study of telehealth encounters from the nationwide HTTT program. The program provided free at-home COVID-19 or influenza test kits, a digital platform for test result submission, and telehealth consultations for clinical evaluation and antiviral prescribing when appropriate. The analytic period spanned the program’s initial pilot implementation in selected communities through nationwide expansion, from September 2023 to July 2024.

### Participants

The initial cohort included 8,160 de-identified individuals who completed at least one HTTT telehealth consultation during the study period. Of these, 1,790 enrolled in an optional survey-based sub-study assessing sociodemographic and digital access characteristics (e.g., internet access, device availability, and digital health literacy). For individuals with multiple encounters, only the first consultation was included to ensure independence of observations.

After excluding participants with missing key demographic variables (n = 745), care-timeline interval data (n = 1,107), or telehealth process duration data (n = 95), the final analytic sample for clustering included 6,213 participants. Among these, 1,023 (16.5%) completed a 10-day follow-up survey assessing symptom trajectories and subsequent healthcare utilization.

### Data Sources and Variables

Telehealth encounter data included consultation modality, defined as the participant’s selected modality preference at the time of the telehealth request, as recorded in the platform. Modality was categorized as synchronous (live video or telephone) or asynchronous (store-and-forward or text-based interactions). This variable reflects participant preference at the time of request; it does not represent the modality ultimately completed or any modality assigned under state-level program requirements. Timing of consultation was categorized as weekday business hours (Monday–Friday, 9:00 AM–5:00 PM, based on the participant’s local time zone) versus after-hours. Provider-documented antiviral prescribing during the encounter (yes/no) was also extracted from the platform.

Care timelines were quantified using two system-recorded intervals (Figure 2): Interval A, the time from positive test result submission to telehealth initiation; and Interval B, the time from telehealth initiation to clinical disposition, defined as the timestamp of the provider’s documented action within the platform (e.g., antiviral prescription issuance or referral to primary care, in-person evaluation, or follow-up services). Interval A may include intermediate platform-based steps occurring between test result submission and telehealth initiation—such as completion of clinical intake surveys, selection of modality preferences, and care logistics (e.g., medication delivery options)—and therefore reflects the full pre-initiation workflow rather than a single discrete transition. For asynchronous encounters, telehealth initiation was defined as the timestamp of the first provider response within the platform. Because the platform did not capture discrete end-of-visit timestamps (e.g., video termination or final message), Interval B was used as a proxy for telehealth process duration following initiation (Figure 2).

**Figure 2.**
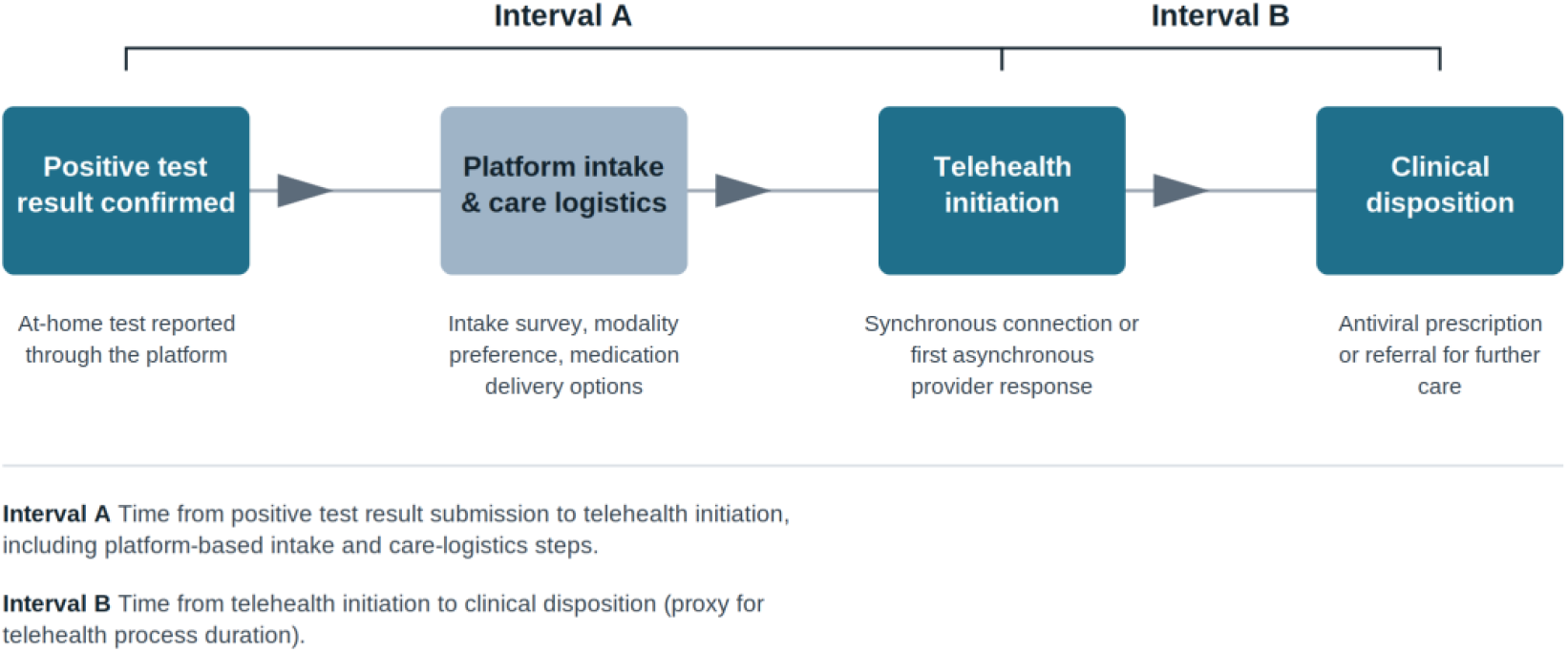
Timeline of key Home Test-to-Treat (HTTT) events and definitions of Intervals A and B. Schematic of the HTTT care process from positive test confirmation to clinical disposition. Interval A denotes the time from positive test result submission to telehealth initiation and encompasses intermediate platform-based steps (clinical intake, modality preference, and care-logistics selections). Interval B denotes the time from telehealth initiation to clinical disposition and serves as a proxy for telehealth process duration. For synchronous encounters, telehealth initiation corresponds to a successful live connection; for asynchronous encounters, it corresponds to the first provider response within the platform.

Survey-derived variables included age, gender, race and ethnicity, insurance status, educational attainment, household income, confidence in the ability to obtain healthcare, reported reasons for delayed healthcare access, and receipt of test kits. The presence of chronic medical conditions (e.g., diabetes, hypertension) was recorded as a binary covariate.

### Outcome Measures

Outcomes were assessed using a self-reported 10-day follow-up survey administered after the index telehealth consultation. Symptom duration was categorized as: never had symptoms; 1–3 days; 4–7 days; 8–10 days; or still symptomatic at follow-up. Subsequent healthcare utilization within 10 days of the index encounter was assessed by participant self-report and included emergency department visits, urgent care visits, additional HTTT telehealth encounters, non-HTTT telehealth visits, and primary care provider visits.

### Statistical Analysis

Telehealth access phenotypes were derived using an unsupervised clustering approach. We performed k-means clustering on standardized (z-score–transformed) variables capturing encounter timing (business hours vs after-hours), modality preference (synchronous vs asynchronous), and telehealth process duration, together with sociodemographic and digital access attributes. A three-cluster solution was selected a priori based on interpretability and separation across candidate solutions. Care-timeline and process measures were included in phenotype derivation. Downstream outcomes, including symptom duration and post-consultation healthcare utilization, were not used in clustering and were analyzed separately to examine associations with access phenotypes.

Descriptive statistics summarized participant characteristics, telehealth utilization measures, care-timeline intervals, and 10-day follow-up outcomes across phenotype groups. Comparisons across groups used analysis of variance for continuous variables and chi-square or Fisher’s exact tests for categorical variables, as appropriate. All analyses were performed in R (version 4.3.3), and statistical significance was defined as a two-sided p-value < 0.05.

### Design Considerations

Analyses were restricted to encounters with complete data for the required timing variables— Interval A (positive test result submission to telehealth initiation) and Interval B (telehealth initiation to clinical disposition)—as well as complete healthcare utilization outcomes. This restriction reduced misclassification and measurement error in care-timeline estimates arising from incomplete timestamp data. Data from synchronous (live video or telephone) and asynchronous (store-and-forward or text-based) encounters were harmonized within a unified analytic dataset to ensure consistent variable definitions and timestamp conventions across modalities.

### Ethics Approval

The study was approved by the University of Massachusetts Chan Medical School Institutional Review Board. All data were de-identified prior to analysis. Participation in the optional research sub-study was voluntary, and written informed consent was obtained from all survey respondents.

## Results

### Telehealth Access Phenotypes

Clustering identified three distinct telehealth access phenotypes among 6,213 participants: Delayed/Disrupted Access (n = 1,537; 24.7%), Digitally Engaged but Socioeconomically Vulnerable (n = 1,460; 23.5%), and Mainstream Access and Efficient Utilization (n = 3,216; 51.8%).

Participants in the Delayed/Disrupted Access group were more likely to initiate encounters outside business hours and had substantially longer telehealth process durations than the other two phenotypes (Interval B mean [SD], 15.93 [3.84] vs 3.69 [3.31] vs 2.87 [2.41] hours, respectively; p < 0.001). This group also reported lower confidence in obtaining healthcare and a higher number of barriers to healthcare access.

The Digitally Engaged but Socioeconomically Vulnerable group was younger, had a higher proportion of female participants, and was more likely to be uninsured, have lower educational attainment, and report lower household income. Participants in this group more frequently selected asynchronous modality preferences, consistent with greater engagement with digital, platform-based care, and more commonly reported financial and access-related barriers.

The Mainstream Access and Efficient Utilization group was characterized by higher rates of insurance coverage, fewer reported barriers to care, greater use of telehealth during business hours, and shorter care-timeline intervals.

### Participant Characteristics and Telehealth Utilization

Across phenotypes, significant differences were observed in age, gender, race and ethnicity, educational attainment, household income, insurance status, prevalence of chronic conditions, confidence in obtaining healthcare, and number of reported reasons for delayed healthcare access (all p < 0.01; Table 1). Synchronous modality preference was lower in the Digitally Engaged but Socioeconomically Vulnerable group (22.9%) than in the Delayed/Disrupted Access (29.8%) and Mainstream Access and Efficient Utilization (30.0%) groups. Despite this lower synchronous preference, the Digitally Engaged but Socioeconomically Vulnerable group had substantially shorter Interval B durations than the Delayed/Disrupted Access group. Antiviral prescription rates were high across all phenotypes (88.6%–91.9%). Mean Interval A and mean Interval B durations differed significantly across phenotypes, with the longest Interval B durations in the Delayed/Disrupted Access group (15.93 hours; p < 0.001; Table 2).

**Table 1.** **Participant characteristics and access vulnerability by phenotype**.

| Characteristic | Delayed/<br>Disrupted<br>Access<br>(n = 1,537) | Digitally Engaged<br>but<br>Socioeconomically<br>Vulnerable (n =<br>1,460) | Mainstream<br>Access<br>and Efficient<br>Utilization (n<br>= 3,216) | P<br>value* |
| --- | --- | --- | --- | --- |
| **Age group, n (%)** |  |  |  | <0.001 |
| 18–45 | 888 (57.8) | 1,077 (73.8) | 1,837 (57.1) |  |
| 46–65 | 438 (28.5) | 269 (18.4) | 959 (29.8) |  |
| 65+ | 211 (13.7) | 114 (7.8) | 420 (13.1) |  |
| **Gender, n (%)** |  |  |  | <0.001 |
| Female | 959 (62.4) | 1,009 (69.1) | 1,985 (61.7) |  |
| Male | 573 (37.3) | 439 (30.1) | 1,224 (38.1) |  |
| Other | 5 (0.3) | 12 (0.8) | 7 (0.2) |  |
| **Race/ethnicity, n (%)** |  |  |  | 0.002 |
| Hispanic | 161 (10.5) | 193 (13.2) | 350 (10.9) |  |
| Non-Hispanic Black | 95 (6.2) | 104 (7.1) | 176 (5.5) |  |
| Non-Hispanic White | 997 (64.9) | 865 (59.2) | 2,115 (65.8) |  |
| Non-Hispanic other | 284 (18.5) | 298 (20.4) | 575 (17.9) |  |
| **Highest education,† n (%)** |  |  |  | <0.001 |
| High school or technical<br>school | 72 (19.9) | 111 (32.2) | 151 (21.1) |  |
| Bachelor's degree | 144 (39.9) | 128 (37.1) | 280 (39.1) |  |
| Master's or doctoral<br>degree | 145 (40.2) | 106 (30.7) | 286 (39.9) |  |
| **Annual family income,† n**<br>**(%)** |  |  |  | <0.001 |
| < \$30,000 | 65 (20.1) | 88 (28.4) | 85 (13.1) | |
| \$30,000 to < \$50,000 | 50 (15.4) | 65 (21.0) | 73 (11.2) | |
| \$50,000 to < \$80,000 | 62 (19.1) | 62 (20.0) | 111 (17.1) | |
| \$80,000 to < \$125,000 | 62 (19.1) | 56 (18.1) | 155 (23.8) | |
| > \$125,000 | 85 (26.2) | 39 (12.6) | 227 (34.9) | |
| Chronic conditions, n (%) | 842 (54.8) | 916 (62.7) | 1,682 (52.3) | <0.001 |
| **Insurance status, n (%)** |  |  |  | <0.001 |
| Employer-provided or<br>self-purchased | 687 (44.7) | 463 (31.7) | 1,785 (55.5) |  |
| Medicaid | 200 (13.0) | 199 (13.6) | 251 (7.8) |  |
| Medicare | 245 (15.9) | 136 (9.3) | 452 (14.1) |  |
| Uninsured (out of<br>pocket) | 303 (19.7) | 557 (38.2) | 486 (15.1) |  |
| Other | 102 (6.6) | 105 (7.2) | 242 (7.5) |  |
| Low/no confidence in<br>obtaining healthcare, n (%) | 959 (62.4) | 1,195 (81.8) | 1,783 (55.4) | <0.001 |
| **No. of reasons delaying**<br>**healthcare access, n (%)** |  |  |  | <0.001 |
| 0 | 639 (41.6) | 40 (2.7) | 1,887 (58.7) |  |
| 1 | 655 (42.6) | 391 (26.8) | 1,329 (41.3) |  |
| 2 | 201 (13.1) | 733 (50.2) | 0 (0.0) |  |
| 3 | 42 (2.7) | 296 (20.3) | 0 (0.0) |  |
| Payment concerns, n (%) | 592 (38.5) | 1,154 (79.0) | 758 (23.6) | <0.001 |
| Limited provider access, n<br>(%) | 237 (15.4) | 718 (49.2) | 263 (8.2) | <0.001 |
| Unable to access healthcare<br>visit,‡ n (%) | 354 (23.0) | 873 (59.8) | 308 (9.6) | <0.001 |

\*P values were calculated using analysis of variance for continuous variables and chi-square tests for categorical variables.
†Education and annual family income were collected only in the research sub-study.
‡Self-reported barrier to accessing healthcare in the prior 12 months (e.g., transportation, inability to take time off work, lack of childcare).

**Table 2.** Telehealth delivery by phenotype.

| Measure | Delayed/<br>Disrupted<br>Access<br>(n = 1,537) | Digitally Engaged<br>but<br>Socioeconomically<br>Vulnerable (n =<br>1,460) | Mainstream<br>Access<br>and Efficient<br>Utilization (n<br>= 3,216) | P<br>value* |
| --- | --- | --- | --- | --- |
| Synchronous modality<br>preference, n (%) | 458 (29.8) | 335 (22.9) | 964 (30.0) | <0.001 |
| Antiviral prescribed by<br>provider, n (%) | 1,362 (88.6) | 1,342 (91.9) | 2,955 (91.9) | <0.001 |
| Requested and received<br>test kits,† n (%) | 229 (14.9) | 338 (23.2) | 422 (13.1) | <0.001 |
| Interval A: test to<br>telehealth initiation (min),<br>mean (SD) | 16.98 (10.25) | 21.27 (17.05) | 13.66 (7.26) | <0.001 |
| Telehealth during business<br>hours, n (%) | 98 (6.4) | 390 (26.7) | 1,030 (32.0) | <0.001 |
| Interval B: initiation to<br>disposition (h), mean<br>(SD)‡ | 15.93 (3.84) | 3.69 (3.31) | 2.87 (2.41) | <0.001 |

\*P values were calculated using analysis of variance for continuous variables and chi-square tests for categorical variables.
†Reflects participants who requested and received HHTT-provided test kits; some participants used personal or externally obtained tests and therefore did not request kits.
‡Interval B is a proxy for telehealth process duration following telehealth initiation.

### Symptoms and Subsequent Healthcare Utilization

Symptom duration did not differ significantly across phenotypes (Table 3). Emergency department visits within 10 days of the index encounter were uncommon overall but more frequent in the Digitally Engaged but Socioeconomically Vulnerable group (2.3%) than in the Delayed/Disrupted Access (0.0%) and Mainstream Access and Efficient Utilization (0.5%) groups (p = 0.02). Urgent care visits were most frequent in the Delayed/Disrupted Access group (5.8%) compared with the Digitally Engaged but Socioeconomically Vulnerable (4.1%) and Mainstream Access and Efficient Utilization (2.0%) groups (p = 0.02). Rates of additional HTTT telehealth encounters, other telehealth visits, and primary care visits did not differ significantly across phenotypes.

**Table 3.**
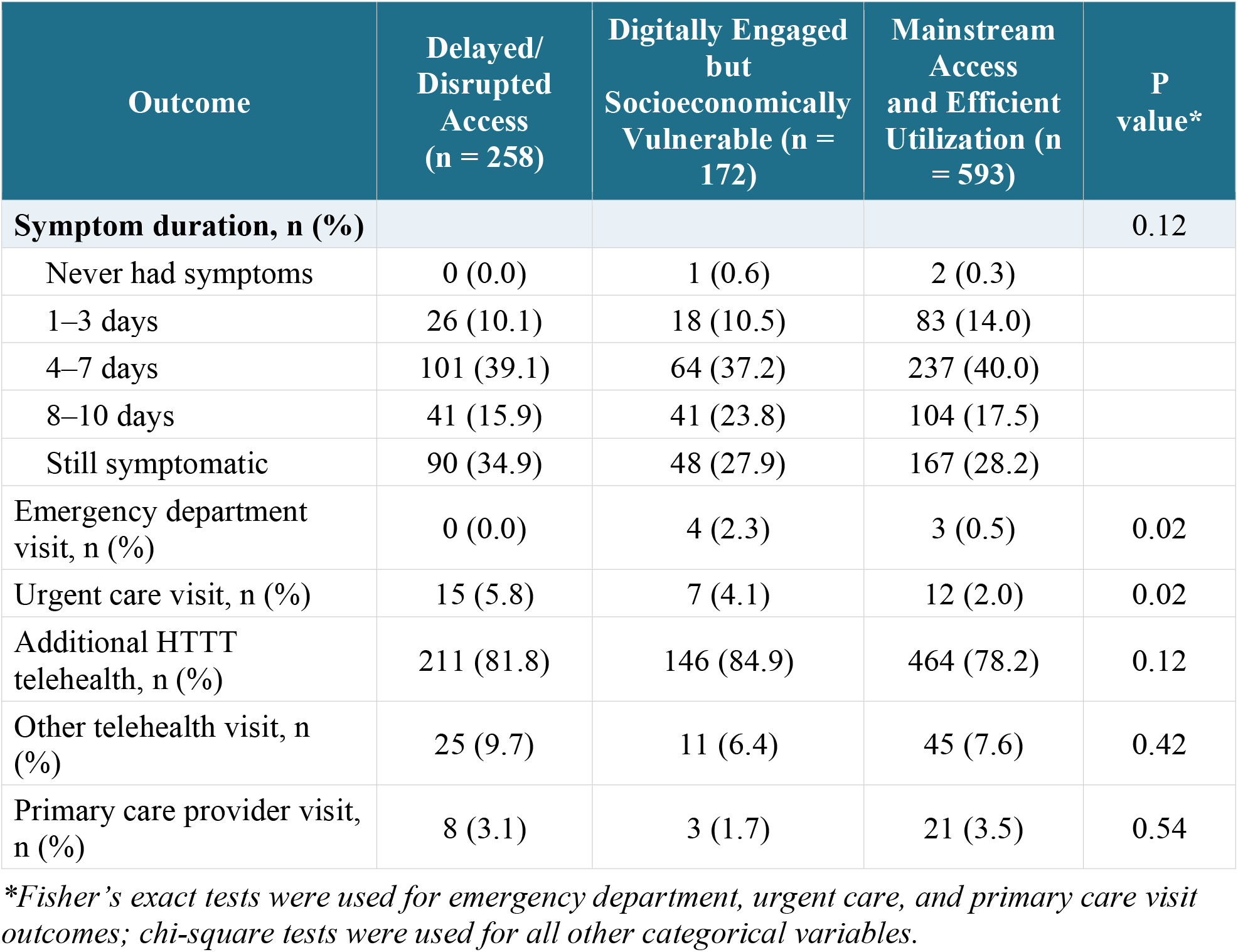
Symptom duration and subsequent healthcare utilization by phenotype, among 10-day follow-up survey respondents.

## Discussion

### Principal Findings

We identified three distinct telehealth access phenotypes: Delayed/Disrupted Access, Digitally Engaged but Socioeconomically Vulnerable, and Mainstream Access and Efficient Utilization. The “digitally engaged” designation reflects a greater preference for asynchronous, platform-based modalities, whereas “socioeconomically vulnerable” reflects a higher burden of uninsurance, lower income and educational attainment, and greater reported access-related barriers. These phenotypes show that telehealth access reflects not only digital engagement but also underlying structural vulnerability and care-seeking behavior, with implications for the design and scaling of home-based testing and treatment programs. Care timelines differed substantially across phenotypes, with the longest durations in the Delayed/Disrupted Access group, intermediate durations in the Digitally Engaged but Socioeconomically Vulnerable group, and the shortest in the Mainstream Access and Efficient Utilization group. These differences highlight heterogeneity in telehealth care processes that visit-based frequency measures alone may not capture. Together, the findings support conceptualizing telehealth access as a heterogeneous process shaped by timing, modality, and care-process efficiency (Figure 2).

### Delayed/Disrupted Access

Participants in this phenotype experienced longer HTTT telehealth process durations, lower confidence in obtaining conventional healthcare, greater reliance on after-hours encounters, and an intermediate burden of reported access barriers—higher than the Mainstream Access and Efficient Utilization group but lower than the Digitally Engaged but Socioeconomically Vulnerable group. These patterns indicate vulnerability in accessing conventional healthcare despite the nominal availability of HTTT telehealth, driven by time scarcity, competing responsibilities, and limited scheduling flexibility, consistent with prior work on structural contributors to health inequities.^12,13^ Such structural and logistical barriers likely contribute to the longer HTTT care timelines observed in this phenotype. These longer durations may also reflect system-level factors, including delays between participant initiation and provider response during after-hours periods, rather than patient-level barriers alone.

Individuals facing inflexible work schedules, caregiving demands, transportation barriers, and limited appointment availability often struggle to engage with conventional healthcare even when services are available.^12–14^ HTTT telehealth may mitigate some of these barriers by reducing travel time and expanding scheduling flexibility,^6,7^ but its effectiveness depends on integration with broader care delivery systems and workflows.^10,15^

For time-sensitive conditions, delays of even a few hours may affect patient experience, trust, and subsequent care-seeking. The higher rate of urgent care utilization in this phenotype likely reflects both acute clinical need and persistent difficulty accessing timely, continuous conventional care. This interpretation aligns with prior work linking structural barriers and time constraints to episodic care^12,13^ and with evidence that patients use telehealth for convenience but still seek additional care—including urgent care or emergency department services—when conventional access remains fragmented.^14^

HTTT programs may function best as complementary access points within a broader care ecosystem rather than stand-alone substitutes for longitudinal conventional care. This view is supported by qualitative interviews with health system leaders highlighting challenges in integrating digital care into existing systems.^15^ Strategies such as targeted after-hours staffing, streamlined asynchronous triage, and proactive navigation support for participants reporting low confidence or multiple access barriers may help reduce delayed or disrupted HTTT access. Telehealth interventions are most likely to advance equity when they are integrated with existing infrastructure, support continuity, and address the structural constraints that limit timely access to conventional care.^10,16–18^

### Digital Engagement and Socioeconomic Vulnerability

Participants in this phenotype more frequently selected asynchronous modality preferences, distinguishing them from the other phenotypes and suggesting a stronger preference for digital, platform-based workflows. Asynchronous telemedicine can offer flexibility and convenience for individuals with competing responsibilities or limited scheduling flexibility, particularly when traditional appointments are difficult to arrange.^19^ Although asynchronous modalities may improve flexibility, underlying socioeconomic barriers likely contribute to the intermediate HTTT care timelines observed in this group. Notably, despite the lowest synchronous modality preference, this phenotype did not have the longest Interval B durations—those were observed in the Delayed/Disrupted Access group. Modality preference alone should therefore not be interpreted as a proxy for delayed provider response or longer telehealth duration.

Despite this digital engagement, these participants had higher rates of uninsurance, lower educational attainment, lower household income, and greater financial and access-related barriers. Digital fluency alone does not ensure equitable access to conventional healthcare; individuals may use digital tools effectively yet remain constrained by broader socioeconomic barriers that limit access to longitudinal care. Higher rates of emergency department utilization may reflect differences in insurance coverage, as emergency departments provide care regardless of insurance status, whereas urgent care and other outpatient settings often require insurance or out-of-pocket payment.^17,18^

These findings have implications for payment and delivery policy. Reimbursement structures and implementation decisions shape access to telehealth, continuity of care, and sustainability.^10,16–18^ Programs such as HTTT are designed to improve access to timely, condition-specific care, particularly for individuals facing barriers to conventional healthcare. However, without durable payment models and integration with existing systems, individuals in this phenotype may continue to rely on episodic telehealth rather than longitudinal primary care, potentially reinforcing fragmentation.

### Mainstream Access and Efficient Utilization

Participants in this phenotype had higher rates of insurance coverage, fewer reported barriers to care, greater use of telehealth during business hours, and shorter HTTT care timelines. This pattern reflects a more routine access pathway in which telehealth is integrated into existing workflows and used under more favorable structural conditions. Rather than representing a normative standard, this phenotype serves as a comparator for understanding how differences in timing, modality, and structural constraints contribute to less efficient access pathways in the Delayed/Disrupted Access and Digitally Engaged but Socioeconomically Vulnerable groups. These structural advantages likely contribute to the shorter HTTT care timelines observed here.

Across phenotypes, symptom duration did not differ significantly, and overall rates of subsequent healthcare utilization were low. These findings suggest that variation in telehealth access processes was not associated with meaningful differences in the short-term (10-day) clinical course or downstream utilization. Prior studies similarly suggest that telehealth implementation does not consistently increase downstream healthcare utilization.^20^

### Strengths and Limitations

A key strength of this study is its process-oriented approach to telehealth access. Rather than relying on visit-based measures, we characterized variation in timing, modality, and care efficiency. This framework may be useful for evaluating equity in large-scale digital health programs designed to deliver rapid, time-sensitive care, by identifying operational delays and structural constraints not captured by utilization measures alone. Phenotype-based analyses may also inform more tailored telehealth delivery strategies, including alignment of synchronous and asynchronous modalities with patient preferences and structural barriers.

Several limitations should be noted. The study was observational and based on routinely collected program and survey data, which limits causal inference. Detailed sociodemographic and digital access variables were available only for participants enrolled in an optional sub-study, introducing potential selection bias and limiting generalizability. Downstream short-term clinical events were infrequent, and differences in subsequent healthcare utilization should be interpreted cautiously. K-means clustering requires prespecification of the number of clusters and may be sensitive to variable selection and scaling, although the resulting phenotypes were conceptually coherent. Finally, system-recorded timestamps do not capture earlier delays related to symptom recognition, testing, or patient decision-making before engagement with the HTTT platform.

### Conclusions

Within a national HTTT program, we identified three telehealth access phenotypes: Delayed/Disrupted Access, Digitally Engaged but Socioeconomically Vulnerable, and Mainstream Access and Efficient Utilization. Telehealth access for time-sensitive conditions can be understood as a process shaped by timing, modality, and structural vulnerability. Evaluating telehealth equity therefore requires attention not only to whether care occurred, but also to how and when it was delivered. A more nuanced understanding of telehealth access processes may help inform the design of future digital health programs aimed at delivering timely, equitable care for acute respiratory infections.

## Acknowledgments

The authors thank the many partners of the Home Test-to-Treat (HTTT) program whose contributions were essential to the program’s implementation but did not meet criteria for authorship. We acknowledge Michael Wolfson, PhD, and Bruce J. Tromberg, PhD (NIH/NIBIB) for conceptual guidance and program leadership; Eric Gogstad (CDC) for expertise in program design and support in providing influenza antivirals; Dina Passman, PhD, MPH, Lisa Tung, PharmD, and Chris Crabtree, DrPH, MPH (ASPR) for contributions to program design and implementation, including the provision of COVID-19 tests and antiviral therapies; Michael Mina, MD, PhD, and the eMed team for adapting the digital platform and implementing the care model; and VentureWell for program coordination and contracting. We also acknowledge the community engagement teams supported by VentureWell for outreach to more than 5,000 community organizations, as well as programmatic support from Alex Guardo, Megan Aanstoos, and Rebekah Neal (VentureWell). We further thank the telehealth clinicians, pharmacists, and staff whose efforts made the HTTT program possible. Most importantly, we are grateful to the HTTT program participants for their trust and participation.

## Author Contributions

W.R.L. conceptualized the study, contributed to the analysis and interpretation of the data, and drafted the manuscript. B.W. contributed to the study design, performed the statistical analysis, and contributed to data interpretation. K.F. and L.O. contributed to data interpretation and critical revision of the manuscript. A.S. contributed to the study design, data acquisition, and interpretation. B.S.G. supervised the work and contributed to the study design, interpretation, and critical revision of the manuscript. All authors reviewed and approved the final manuscript and agree to be accountable for all aspects of the work.

## Statements and Declarations

### Ethical Considerations

The study was approved by the University of Massachusetts Chan Medical School Institutional Review Board. All data were de-identified prior to analysis. The study was conducted in accordance with the principles of the Declaration of Helsinki.

### Consent to Participate

Participation in the optional research sub-study was voluntary, and written informed consent was obtained from all survey respondents.

### Consent for Publication

Not applicable. The manuscript does not contain any individual person’s identifiable data.

### Data Availability Statement

The de-identified data that support the findings of this study may be made available from the corresponding author upon reasonable request, subject to the data-use agreements and privacy protections governing the Home Test-to-Treat program.

